# Separating the direct effects of risk factors for atherosclerotic cardiovascular disease from those mediated by type 2 diabetes

**DOI:** 10.1101/2021.08.05.21261658

**Authors:** Venexia M Walker, Marijana Vujkovic, Alice R Carter, Neil M Davies, Miriam S Udler, Michael G Levin, George Davey Smith, Benjamin F Voight, Tom R Gaunt, Scott M Damrauer

## Abstract

**Background:** Type 2 diabetes and atherosclerotic cardiovascular disease share several risk factors. However, it is unclear whether the effect of these risk factors on liability to atherosclerotic cardiovascular disease is independent of their effect on liability to type 2 diabetes.

**Methods:** We performed univariate Mendelian randomization to quantify the effects of continuous risk factors from the IEU OpenGWAS database on liability to three outcomes: type 2 diabetes, coronary artery disease, and peripheral artery disease, as well as the effects of liability to type 2 diabetes on the risk factors. We also performed two-step Mendelian randomization for mediation to estimate the mediating pathways between the risk factors, liability to type 2 diabetes, and liability to the atherosclerotic cardiovascular disease outcomes where possible.

**Results:** We found evidence for 53 risk factors as causes of liability to coronary artery disease, including eight which were causes of liability to type 2 diabetes only and four which were consequences only. Except for fasting insulin and hip circumference, the direct and total effects from the two-step Mendelian randomization were similar. This suggests that the combination of these risk factors with liability to type 2 diabetes was unlikely to alter liability to coronary artery disease beyond their individual effects. We also found 13 risk factors that were causes of liability peripheral artery disease, including six which were causes of liability to type 2 diabetes only and four which were consequences only. Again, the direct and total effects were similar for these ten risk factors apart from fasting insulin.

**Conclusions:** Most risk factors were likely to affect liability to atherosclerotic cardiovascular disease independently of their relationship with liability to type 2 diabetes. Control of modifiable risk factors therefore remains important for reducing atherosclerotic cardiovascular disease risk regardless of patient liability to type 2 diabetes.

**RESEARCH IN CONTEXT:** *What is already known about this subject?:*

- Type 2 diabetes, coronary artery disease and peripheral artery disease, share several risk factors
- Type 2 diabetes is also one of the strongest independent risk factors for both coronary and peripheral artery disease

*What is the key question?:*

- Which risk factors for atherosclerotic cardiovascular disease are mediated by liability to type 2 diabetes and which are independent?

*What are the new findings?:*

- Among 108 risk factors in this study, there was evidence to support: 10 risk factors as causes, 23 risk factors as consequences, and 34 risk factors as both causes and consequences of liability to type 2 diabetes
- In addition, we found evidence for 53 risk factors as causes of liability to coronary artery disease and 42 risk factors as causes of liability to peripheral artery disease
- Using two-step Mendelian randomization for mediation, we found most risk factors for atherosclerotic cardiovascular disease were likely to act independently of liability to type 2 diabetes

*How might this impact on clinical practice in the foreseeable future?:*

- Our findings support continued control of modifiable risk factors as this is likely to reduce atherosclerotic cardiovascular disease, regardless of patient liability to type 2 diabetes

## INTRODUCTION

Type 2 diabetes and atherosclerotic cardiovascular diseases, coronary artery disease and peripheral artery disease, share several risk factors, such as obesity and hypertension (1–3). In addition, type 2 diabetes is one of the strongest independent risk factors for both coronary and peripheral artery disease (4,5). Primary prevention strategies for both diabetes and atherosclerotic cardiovascular disease focus on lifestyle modification to improve the shared set of cardiometabolic risk factors including obesity, hypertension, and dyslipidaemia. (6,7) However, despite the shared links between cardiometabolic risk factors, diabetes, and atherosclerotic cardiovascular disease, the effects of risk factors on liability to atherosclerotic cardiovascular disease, both through and independent of liability to type 2 diabetes, has not been systematically assessed.

Mendelian randomization uses genetic variants associated with an exposure – referred to as an ‘instrument’ – as a proxy for that exposure. (8) Mendelian randomization can be used to estimate the causal effect of an exposure on an outcome free from bias due to non-genetic confounding and reverse causality if its assumptions hold (**Supplementary Text 1**). (9) Two-step Mendelian randomization for mediation analysis is an extension to this method, which incorporates the causal effect of a mediator, to estimate the direct (independent of the mediator) and indirect (via the mediator) effects of an exposure on an outcome. (10,11) Furthermore, this approach can be applied using summary statistics from multiple genome-wide association studies (GWASs) with non-overlapping samples. (12) This removes the need for individual level data from a single study containing information on all the risk factors.

Mendelian randomization has previously been used to individually estimate the effect of several risk factors on liability to our three disease outcomes of interest: type 2 diabetes, coronary artery disease, and peripheral artery disease. (4,13–17) Mendelian randomization for mediation has also been conducted to investigate the effect of a selected set of obesity-related markers (genetically predicted BMI and waist-hip ratio) on liability to coronary artery disease, peripheral artery disease and stroke, mediated by genetically predicted systolic blood pressure, liability to type 2 diabetes, lipid risk factors and smoking. (18) However, systematic assessment of a wide range of risk factors using Mendelian randomization to separate their effects on liability to atherosclerotic cardiovascular disease from liability to type 2 diabetes has not yet been conducted.

In this study, we implemented a standardized univariate Mendelian randomization framework and follow-up analyses with two-step Mendelian randomization for mediation to interrogate the association of a broad range of continuous risk factors with liability to our three disease outcomes: type 2 diabetes, coronary artery disease, and peripheral artery disease. The aim of this study was to separate the direct effects of the risk factors on atherosclerotic cardiovascular disease from those mediated by liability to type 2 diabetes.

## METHODS

### Study design

Our study consisted of two stages, which are summarised in **Figure 1**. First, we used univariate Mendelian randomization to estimate the effects of 108 continuous risk factors (see: **Risk factor selection**) on liability to three disease outcomes: type 2 diabetes, coronary artery disease, and peripheral artery disease. In addition, we used univariate Mendelian randomization to estimate the effect of liability to type 2 diabetes on the 108 continuous risk factors. This allowed us to remove risk factors that had a bidirectional association with liability to type 2 diabetes, which may indicate interaction of the phenotypes and violate the assumptions required for the subsequent mediation stage. Based on the evidence from stage 1, we implemented stage 2: two-step Mendelian randomization for mediation, which assumes no interaction between the exposure and the mediator. Using this approach, we estimated the direct (i.e., the effect independent of liability to type 2 diabetes) and indirect effects (i.e., the effect mediated via liability to type 2 diabetes) of the risk factors on the atherosclerotic cardiovascular diseases of interest.

**Figure 1:**
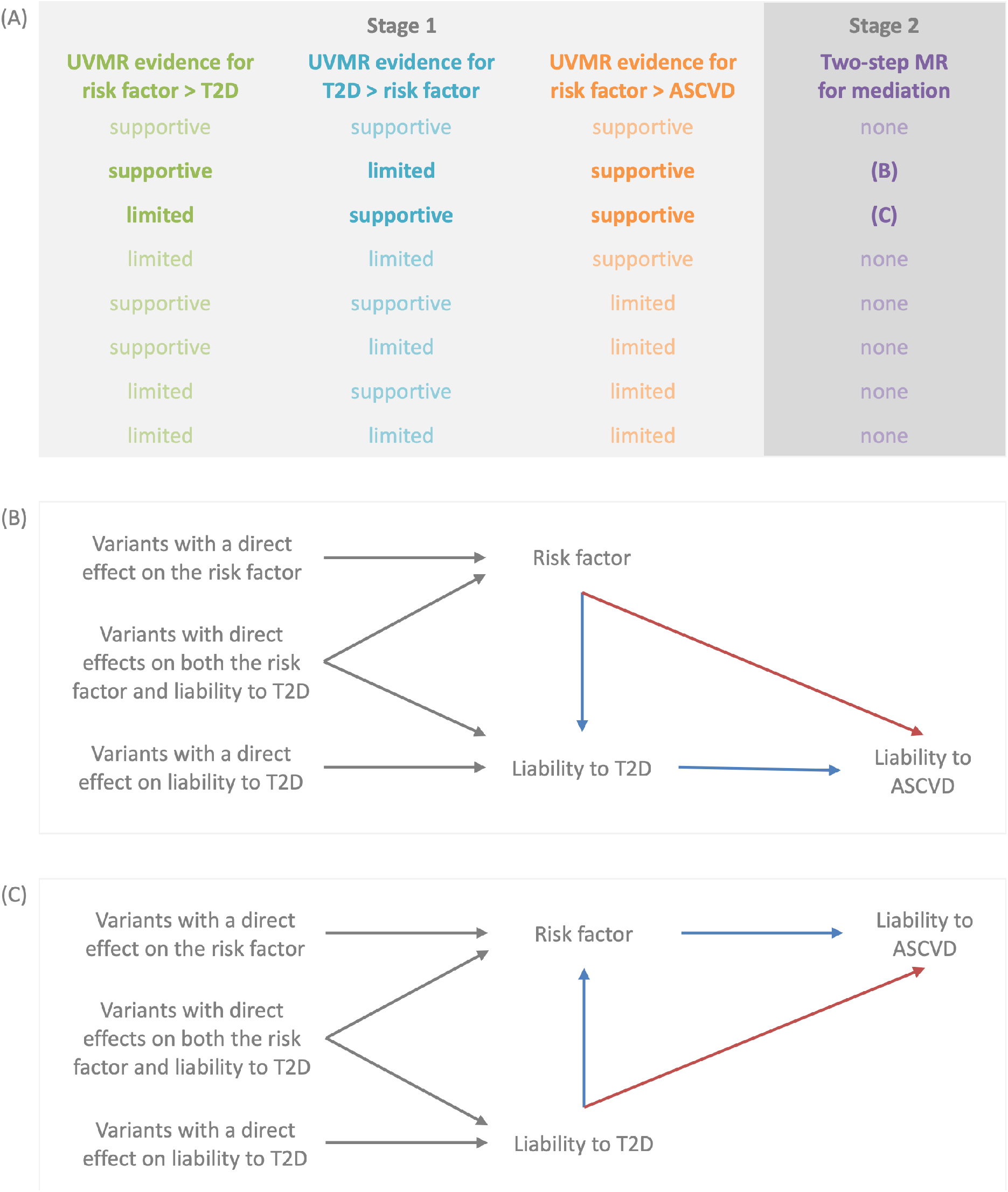
Illustration of the two-stage study design (panel A) and the two-step Mendelian randomization for mediation models used for stage 2 (panels B and C) Panel A summarizes how evidence from univariate Mendelian randomization (UVMR) analyses of the risk factor, liability to type 2 diabetes (T2D), and liability to atherosclerotic cardiovascular diseases (ASCVD) is assessed in stage 1. Here, estimates that met the arbitrary false discovery rate threshold of 5% were deemed to have ‘supportive’ evidence, while all other estimates were deemed to have ‘limited’ evidence. Stage 1 then informs the analysis models used in stage 2 for the two-step Mendelian randomization (MR) for mediation. These models are described in panels B and C, where the red arrows represent the direct (i.e. independent of the mediator) effect and the blue arrows represent the indirect (i.e. via the mediator) effect.

### Risk factor selection

Risk factors were selected from the IEU OpenGWAS database by implementing a selection procedure to retain the largest, minimally adjusted GWAS for each continuous biological trait that had been studied in both men and women of European or mixed ancestry (**Supplementary Figure 1**). (19) Sample overlap was permitted between risk factors and so the majority of GWAS included participants from UK Biobank. (20)

### Outcome phenotypes

We obtained the GWAS for liability to type 2 diabetes in European ancestry from the DIAMANTE consortium. (21) The GWAS for liability to coronary artery disease and liability to peripheral artery disease were obtained from the CARDIoGRAM consortium and Million Veteran Program respectively. (22–24) As noted above, sample overlap was permitted between risk factors, however GWAS were obtained from distinct samples for liability to type 2 diabetes, coronary artery disease, and peripheral artery disease.

### Univariate Mendelian randomization

Instruments for each risk factor were defined using the genome-wide significant (p<5e-8) genetic variants from the corresponding GWAS to satisfy the first instrumental variable assumption of relevance. For the univariate Mendelian randomization analyses, instruments were clumped using a 10Mb window and R squared linkage disequilibrium (LD) threshold of 0.001 against the 1000 genomes reference panel for the European super-population, which was filtered to include only bi-allelic variants with minor allele frequencies greater than 0.01. Instruments consisting of less than 10 variants were removed, before harmonization with the outcome data to represent an increase in the exposure. Mendelian randomization was then performed using the inverse variance weighted method.

We repeated the above univariate Mendelian randomization analyses using the simple mode, weighted median, weighted mode, and MR-Egger methods as a sensitivity analysis to examine estimate consistency. We also derived heterogeneity statistics to examine the consistency of estimates across the variants included in each analysis and performed a leave-one-out analysis to determine whether certain variants were driving the observed effects. We included an MR-Egger intercept test to assess if directional pleiotropy was likely to have affected our results. (25) Finally, to assess the no measurement error assumption for MR-Egger, we calculated the 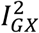 statistic as a measure of potential attenuation bias. (26) All univariate analyses and associated sensitivity analyses were implemented using the TwoSampleMR package for R. (27)

### Two-step Mendelian randomization for mediation

For risk factors with both (i) evidence of a unidirectional association with liability to type 2 diabetes (in either direction) and (ii) evidence of an effect on liability to at least one atherosclerotic cardiovascular outcome of interest, multivariable Mendelian randomization was applied using the risk factor and liability to type 2 diabetes as exposures. As noted previously, evidence of a unidirectional association with liability to type 2 diabetes is necessary, as two-step Mendelian randomization for mediation assumes no interaction between the exposure and the mediator, which we cannot rule out when a bidirectional association between a risk factor and liability to type 2 diabetes is observed. An arbitrary false discovery rate (FDR) of 5%, calculated according to the Benjamini and Hochberg method, was used as an indicator of supportive evidence of an association between risk factors and liability to type 2 diabetes. (28)

Based on the direction of the effect between the risk factor and liability to type 2 diabetes, multivariable Mendelian randomization allowed us to estimate either (i) the effect of the risk factor, independent of liability to type 2 diabetes, on the liability to atherosclerotic cardiovascular outcome of interest (**Figure 1, panel B**) or (ii) the effect of liability to type 2 diabetes, independent of the risk factor, on the liability to atherosclerotic cardiovascular outcome of interest (**Figure 1, panel C**). These effects are often referred to as ‘direct’ effects. Where appropriate, we also derived either (i) the effect of the risk factor, through liability to type 2 diabetes, or (ii) the effect of liability to type 2 diabetes, through the risk factor, on liability to the atherosclerotic cardiovascular outcome of interest. These effects are often referred to as ‘indirect’ or ‘mediated’ effects. For these analyses, we multiplied the estimate for the effect of the exposure of interest on the mediator obtained from the univariate Mendelian randomization by the direct effect of mediator on the outcome obtained from the multivariable Mendelian randomization (where the exposure of interest and mediator were both used as exposures). Confidence intervals were derived using the sum of squares method.

Instruments for this analysis were clumped against either the risk factor or liability to type 2 diabetes (whichever GWAS had the smallest instrument) using a 10Mb window and R squared LD threshold of 0.001 against the 1000 genomes reference panel for the European super-population, which was filtered to include only bi-allelic variants with minor allele frequencies greater than 0.01. Harmonisation was performed with variants aligned to represent an increase in the exposure prior to analysis. We calculated conditional F statistics to test instrument strength for each exposure in our analysis. We also calculated a modified form of Cochran’s Q statistic that has been developed to measure heterogeneity in causal effect estimates from multivariable Mendelian randomization. Multivariable Mendelian randomization estimates and these statistics were obtained using the MVMR package for R. (29) The non-collapsibility of odds ratios can pose a problem when using summary statistics from logistic regression for binary mediators and outcomes in multivariable Mendelian randomization. To assess whether this is likely to have impacted our results, we repeated our analyses using a GWAS of liability to type 2 diabetes based on a linear (instead of a logistic) model (**Supplementary Text 2**).

### Code availability

All analyses were conducted in R version 4.0.2. The associated code is available from: https://github.com/venexia/T2DMediationMR.

## RESULTS

The results of this analysis are presented in four parts. First, the selection of risk factors from the IEU OpenGWAS database. (19) Second, the results of the univariate Mendelian randomization analyses to interrogate the effect of each risk factor on liability to type 2 diabetes and the effect of liability to type 2 diabetes on each risk factor. Third, the results related to liability to coronary artery disease from both the univariate Mendelian randomization and two-step Mendelian randomization for mediation. Fourth, and finally, results related to liability to peripheral artery disease from the univariate Mendelian randomization and two-step Mendelian randomization for mediation.

### Risk factor selection

We identified 108 risk factors from the IEU OpenGWAS database for inclusion in our analysis. (19) Details of both the risk factor and outcome GWASs are provided in **Supplementary Table 1**. The majority of the risk factor GWASs were conducted in UK Biobank by the Neale lab. (30) Twelve of the selected GWASs were from other sources: adiponectin (31); alcoholic drinks per week (32); body fat (33); body mass index (34); cigarettes per day (32); fasting glucose (35); fasting insulin (35); heart rate (36); neuroticism (37); total cholesterol (38); urinary sodium-potassium ratio (39); and waist-to-hip ratio (40).

### Causes and consequences of liability to type 2 diabetes

Estimates from bidirectional univariate Mendelian randomization of each risk factor and liability to type 2 diabetes found evidence for: 10 risk factors as causes, 23 risk factors as consequences, 34 risk factors as both causes and consequences, and 41 risk factors as neither causes nor consequences of liability to type 2 diabetes at an FDR threshold of 5% (**Supplementary Figures 2 and 3; Supplementary Table 2**). Sensitivity analyses using alternative Mendelian randomization methods were consistent with the inverse variance weighted estimates **(Supplementary Figures 4 and 5; Supplementary Table 2**). The MR-Egger intercept test found intercepts between -0.15 (body fat on liability to type 2 diabetes) and 0.07 (fasting glucose on liability to type 2 diabetes) (**Supplementary Table 3**). Finally, the 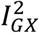 statistic was over 0.93 for all MR Egger results (**Supplementary Table 4**). When taken as an estimate of the attenuation bias in these analyses, this corresponds to less than 7% relative bias towards the null. Based on the findings from these bidirectional univariate Mendelian randomization analyses, 33 of the 108 risk factors were eligible for the two-step Mendelian randomization for mediation analyses for liability to atherosclerotic cardiovascular disease that followed.

### Causes of liability to coronary artery disease

Using univariate Mendelian randomization, we found evidence for 53 of the 108 risk factors as causes of liability to coronary artery disease at the FDR threshold of 5%. Twelve of these risk factors also had a unidirectional association with liability to type 2 diabetes and so were eligible for two-step Mendelian randomization for mediation (**Supplementary Figures 6 and 7; Supplementary Tables 2 and 5**). Using the risk factor as the exposure and liability to type 2 diabetes as the mediator for the eight risk factors identified as causes of both liabilities to type 2 diabetes and coronary artery disease, we found similar direct and total effects for most risk factors: Apolipoprotein B, aspartate aminotransferase, diastolic blood pressure, standing height, total cholesterol, and trunk fat percentage (**Figure 2; Supplementary Table 6**). The exceptions were fasting insulin and hip circumference, where the effects indicated partial mediation by liability to type 2 diabetes. For the remaining four risk factors (mean corpuscular haemoglobin, mean sphered cell volume, testosterone, urate), we used liability to type 2 diabetes as the exposure and the risk factor as the mediator as they had been identified as consequences, rather than causes, of liability to type 2 diabetes. In this analysis, we found similar direct and total effects in all cases, suggesting that the risk factor and liability to type 2 diabetes were likely to have independent effects on liability to coronary artery disease (**Figure 3; Supplementary Table 6**). The conditional F statistics for the multivariable Mendelian randomization component of these analyses ranged from 9 to 87 (**Supplementary Table 6**), indicating good instrument strength. Meanwhile, the modified Cochran’s Q statistic exceeded the critical value for the Χ^2^ distribution at the 5% level for all analyses. This indicated that the chosen SNPs predicted both the risk factor and liability to type 2 diabetes in the data. Taken as a whole, the analyses concerning liability to coronary artery disease suggest that the effects of the risk factors are likely independent of the effects of liability to type 2 diabetes for the most part.

**Figure 2:**
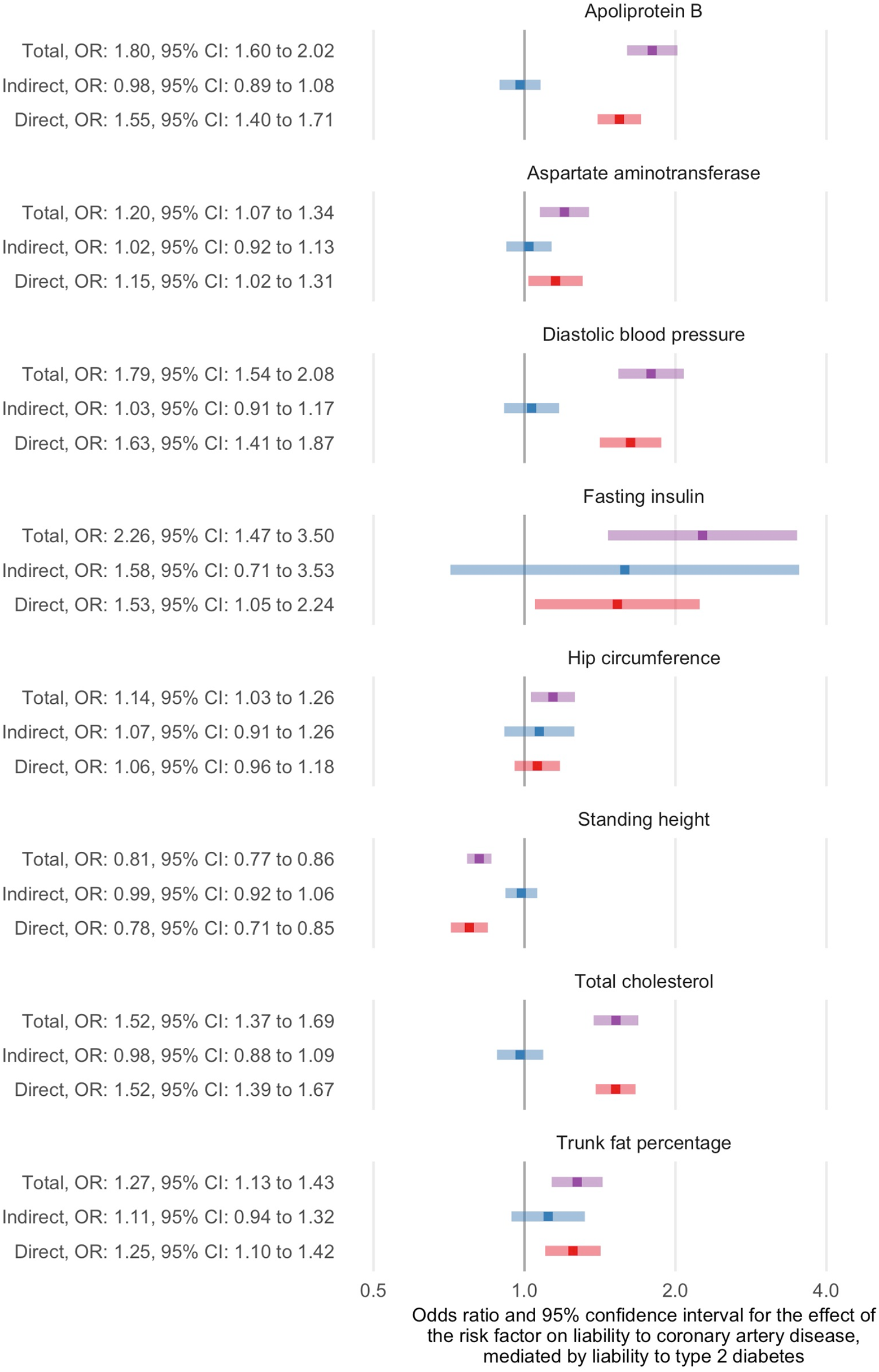
Two-step Mendelian randomization for mediation estimates for the total, indirect (mediated by liability to type 2 diabetes) and direct (independent of liability to type 2 diabetes) effects of the indicated risk factors on liability to coronary artery disease

**Figure 3:**
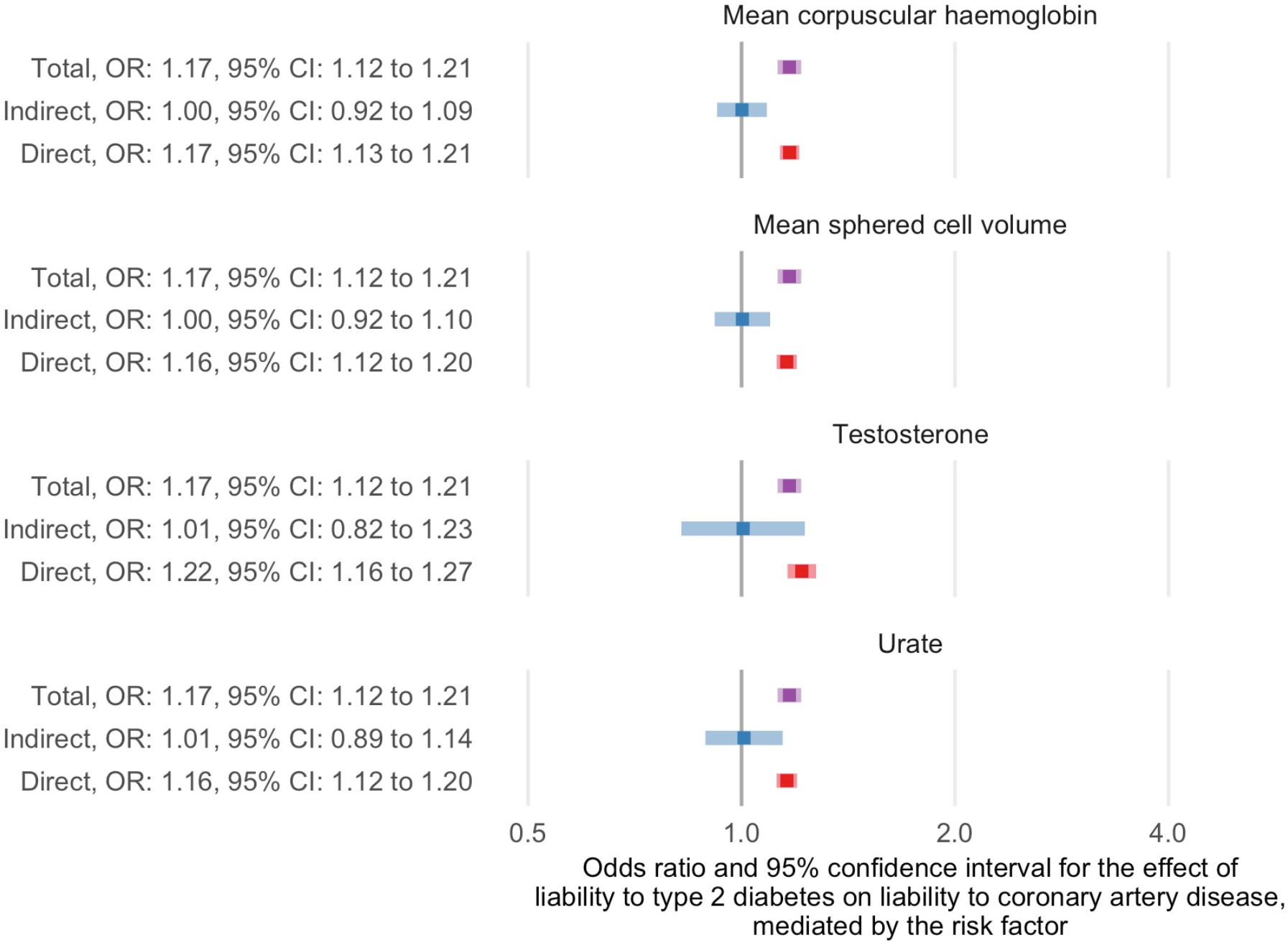
Two-step Mendelian randomization for mediation estimates for the total, indirect (mediated by the indicated risk factor) and direct (independent of the indicated risk factor) effects of liability to type 2 diabetes on liability to coronary artery disease

### Causes of liability to peripheral artery disease

We found evidence for 42 risk factors as causes of liability to peripheral artery disease at the FDR threshold of 5% using univariate Mendelian randomization. We performed two-step Mendelian randomization for mediation on 10 of these 42 risk factors with evidence to support a unidirectional association with liability to type 2 diabetes (**Supplementary Figures 8 and 9; Supplementary Tables 2 and 5**). Using the risk factor as the exposure and liability to type 2 diabetes as the mediator for the six risk factors (Apolipoprotein B, diastolic blood pressure, fasting insulin, hip circumference, total cholesterol, and trunk fat percentage) identified as causes of both liability to type 2 diabetes and liability to coronary artery disease, we found similar direct and total effects in most cases (**Figure 4; Supplementary Table 6**). Fasting insulin was again identified as an exception with effects that indicated partial mediation by liability to type 2 diabetes. For the other four risk factors (alcohol intake frequency, cigarettes per day, sodium in urine, and urate), we used liability to type 2 diabetes as the exposure and the risk factor as the mediator and found similar direct and total effects in all cases (**Figure 5; Supplementary Table 6**). The conditional F statistics for the multivariable Mendelian randomization component of these analyses again indicated good instrument strength, ranging from 9 to 86 (**Supplementary Table 6**). We also found the modified Cochran’s Q statistic exceeded the critical value for the Χ^2^ distribution at the 5% level for all liability to peripheral artery disease analyses. Similar to the results concerning liability to coronary artery disease, these analyses suggest that the effects for the majority of risk factors on liability to peripheral artery disease are likely to be independent of the effects of liability to type 2 diabetes.

**Figure 4:**
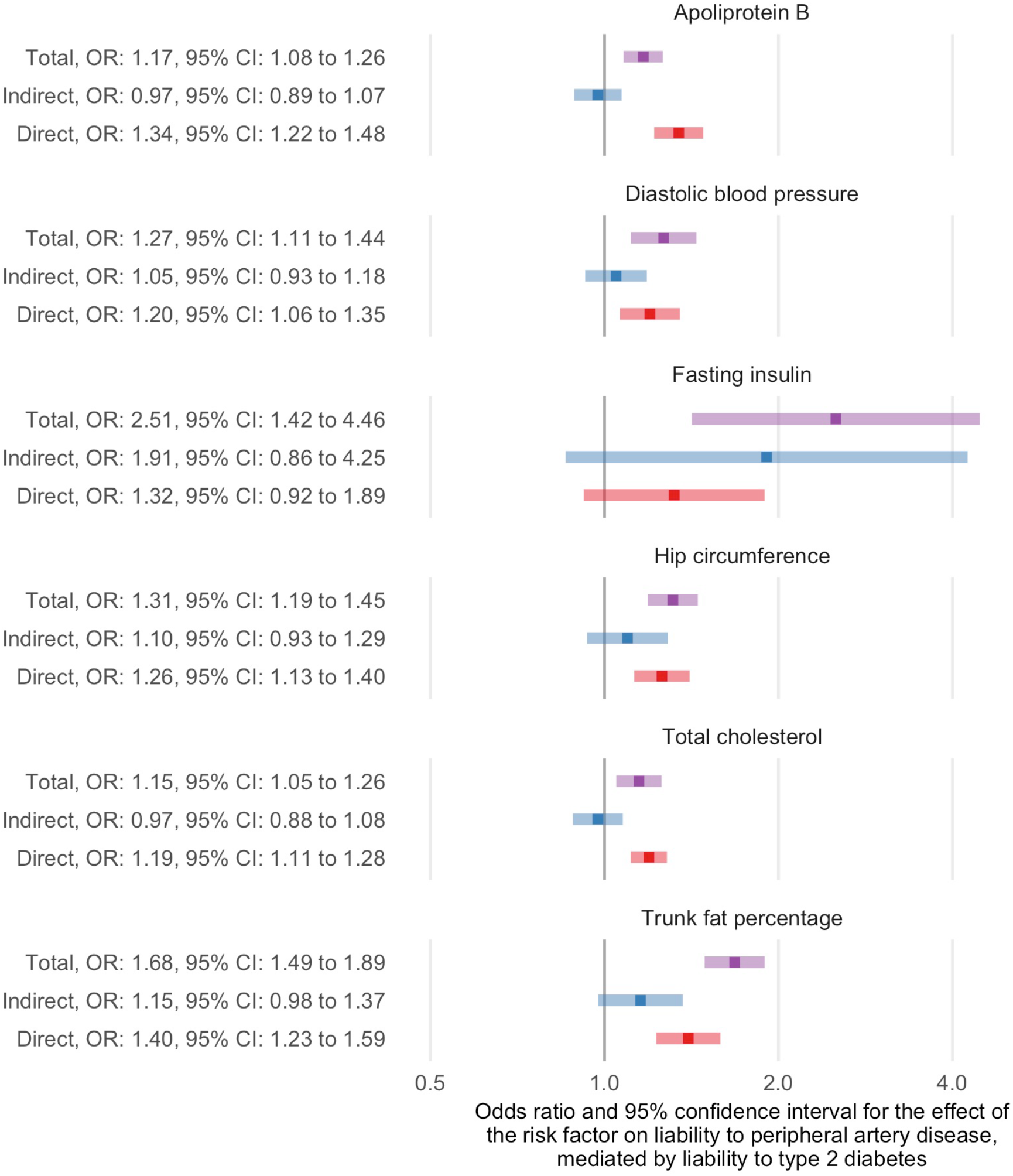
Two-step Mendelian randomization for mediation estimates for the total, indirect (mediated by liability to type 2 diabetes) and direct (independent of liability to type 2 diabetes) effects of the indicated risk factors on liability to peripheral artery disease

**Figure 5:**
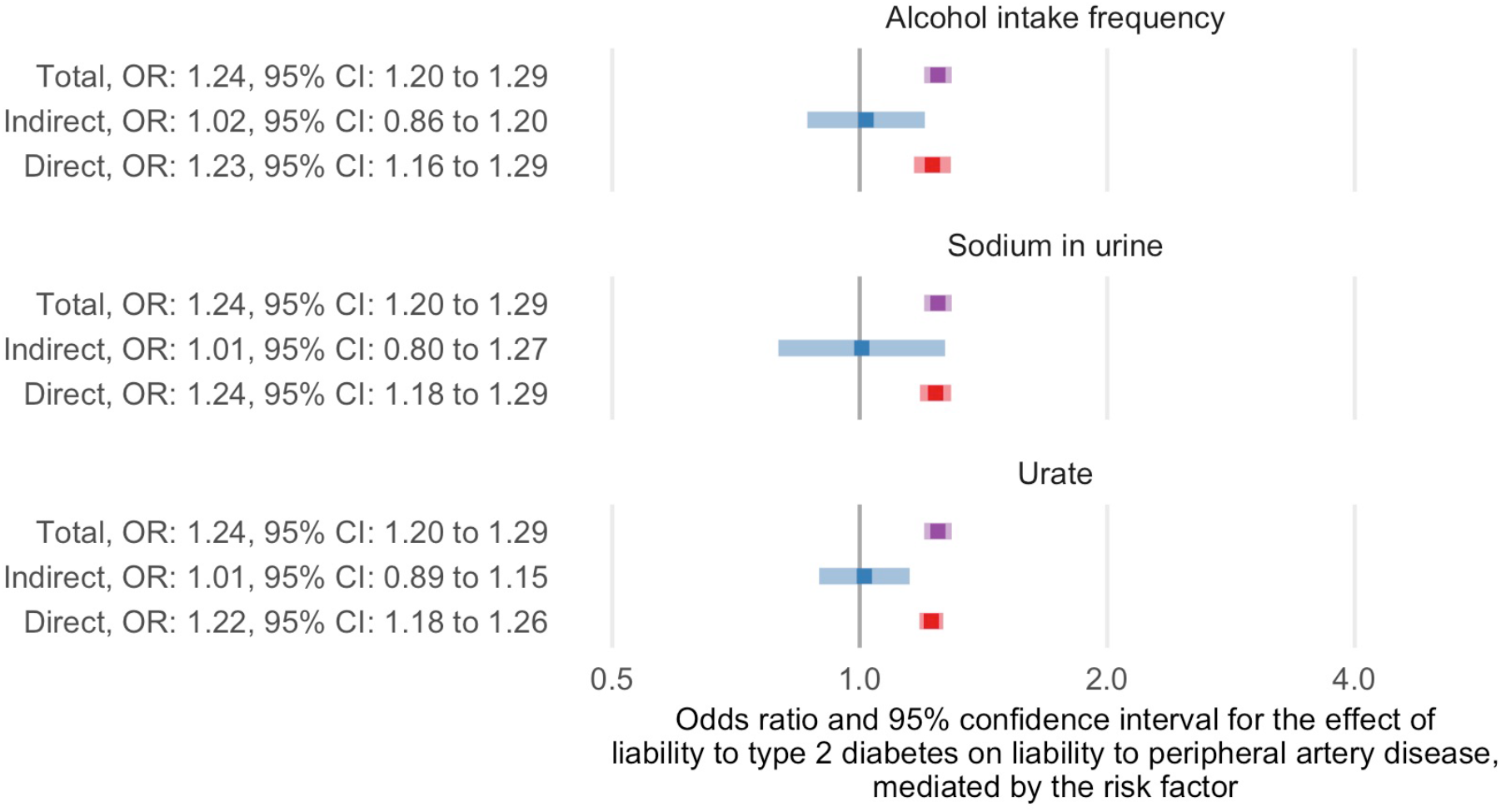
Two-step Mendelian randomization for mediation estimates for the total, indirect (mediated by the indicated risk factor) and direct (independent of the indicated risk factor effects of liability to type 2 diabetes on liability to peripheral artery disease

## DISCUSSION

This study found evidence for the causal effects of multiple risk factors on liability to our three outcomes of interest: type 2 diabetes, coronary artery disease, and peripheral artery disease, using univariate Mendelian randomization. Common risk factors for liability to these outcomes included glycaemic traits, such as glucose (type 2 diabetes: OR = 3.34, 95% CI = 2.41 to 4.63; coronary artery disease, OR = 1.25, 95% CI = 1.11 to 1.41; peripheral artery disease: OR = 1.26, 95% CI = 1.10 to 1.44) and anthropometric traits, such as body fat percentage (type 2 diabetes: OR = 2.78, 95% CI = 2.32 to 3.32; coronary artery disease: OR = 1.52, 95% CI = 1.33 to 1.73; peripheral artery disease: OR = 1.92, 95% CI = 1.68 to 2.19). We also identified specific risk factors for each outcome. For instance, there were five risk factors with evidence to support an effect on liability to type 2 diabetes (whole body fat-free mass, whole body water mass, peak expiratory flow, lymphocyte count, IGF-1) but not liability to coronary or peripheral artery disease; as well as twelve and eight risk factors with specific effects on liability to coronary and peripheral artery disease respectively. These findings support continued control of the modifiable risk factors studied, independent of the diabetes control, to reduce the risk of these outcomes.

Using two-step Mendelian randomization for mediation analysis, this study found that the effects of most of the eligible risk factors were likely to be independent of the effects of liability to type 2 diabetes. There are several reasons why a mediating effect may not have been identified in this analysis. Firstly, there could be no true mediating effect, so our findings reflect reality. Secondly, we may lack power to detect a mediating effect as the power requirements for multivariable Mendelian randomization are greater than univariate approaches and the number of risk factors considered in this study comes with a high multiple testing burden. Alternatively, the phenotypic complexity of liability to type 2 diabetes may be obscuring effects if, for example, a risk factor acts on a certain component of liability to type 2 diabetes that does not have a causal effect on liability to atherosclerotic cardiovascular disease. Partial mediation was observed for two risk factors: fasting insulin, which is difficult to separate from the clinical definition of type 2 diabetes, and hip circumference, though this particular risk factor was only an exception for the outcome liability to coronary artery disease. Several of the risk factors tested, including body mass index and waist-hip-ratio, were identified as both causes and consequences of liability to type 2 diabetes and so were not studied using two-step Mendelian randomization for mediation, even if the magnitude of the effects heavily favoured a direction. Despite this, the strong causal effects observed for these risk factors on liability to coronary and peripheral artery disease, without consideration of liability to type 2 diabetes, indicate that they remain important risk factors for reducing the risk of atherosclerotic cardiovascular disease outcomes.

Four risk factors included in this study may be considered as part of the clinical definition of type 2 diabetes: fasting glucose, fasting insulin, glucose, and glycated haemoglobin (HbA1c). Except for fasting insulin, which was found to be a cause but not a consequence of liability to type 2 diabetes, these risk factors were deemed to have bidirectional relationships with liability to type 2 diabetes when interpreted using the arbitrary 5% FDR threshold selected for this study. Given the interrelated nature of these glycaemic risk factors with liability to type 2 diabetes, this is perhaps unsurprising and highlights the difficultly in disentangling these effects. Nonetheless, these risk factors were important to include in our analysis given our aim of systematically assessing the effects of risk factors on liability to atherosclerotic cardiovascular disease risk.

Our study has some limitations. Mendelian randomization requires several assumptions to hold for valid estimates to be obtained. (8,9) Except for relevance, these assumptions cannot be tested. However, where possible, we have performed sensitivity analyses and falsification tests. A further concern is the assumption of no interaction between risk factors and liability to type 2 diabetes that is necessary for the two-step Mendelian randomization for mediation. We cannot readily test this assumption using summary data, so the impact of any violation is difficult to quantify. For this reason, we were conservative in our approach to the analysis and did not perform two-step Mendelian randomization for mediation on any risk factors that had a bidirectional relationship with liability to type 2 diabetes at the 5% FDR threshold. This should help to minimize the possibility of model misspecification. Our study may also be biased due to the non-collapsibility of odd ratios, which can impact estimates as a result of summary statistics from logistic regression being used for binary mediators (such as liability to type 2 diabetes) and outcomes (such as liability to coronary and peripheral artery disease). (11) We assessed this possibility by repeating our analyses with summary statistics from a novel GWAS that used a linear model for liability to type 2 diabetes and found little difference in the Mendelian randomization estimates we obtained (**Supplementary Text 2; Supplementary Figure 10**). This indicates that non-collapsibility of odds ratio is unlikely to have impacted our results. In addition, our study may be affected by horizontal pleiotropy. We used MR-Egger estimators to investigate whether our results were sensitive to assumptions about the structure of pleiotropy and found some evidence that a small number of risk factors may have horizontally pleiotropic effects. Finally, our study was restricted to individuals of European or mixed ancestry due to the broad range of GWAS required for the analysis. Consequently, the generalizability of the findings from this study is limited to comparable European or mixed ancestry populations.

In conclusion, we have used a Mendelian randomization framework to separate the effects of continuous risk factors from liability to type 2 diabetes and aid our understanding of their relationships with liability to coronary and peripheral artery disease. Our analysis suggests that some key risk factors – including diastolic blood pressure and hip circumference – act independently of liability to type 2 diabetes. Therefore, control of the modifiable risk factors included in this study is likely to reduce atherosclerotic cardiovascular disease, regardless of patient liability to type 2 diabetes.

## Data Availability

All data used in this study are publicly available. We accessed genome-wide association study summary statistics for the risk factors from the IEU OpenGWAS database: https://gwas.mrcieu.ac.uk/; for liability to type 2 diabetes from the DIAMANTE consortium: https://www.diagram-consortium.org/; for liability to coronary artery disease from the CARDIoGRAM consortium: http://www.cardiogramplusc4d.org/; and for liability to peripheral artery disease from dbGAP: https://www.ncbi.nlm.nih.gov/gap/. Approved dbGAP access to phs001672.v6.p1 was provided to B.F.V. (dbGAP project ID: 27398). The code for this study has been made available on GitHub: https://github.com/venexia/T2DMediationMR.

## DATA AVAILABILITY STATEMENT

All data used in this study are publicly available. We accessed genome-wide association study summary statistics for the risk factors from the IEU OpenGWAS database: https://gwas.mrcieu.ac.uk/; for liability to type 2 diabetes from the DIAMANTE consortium: https://www.diagram-consortium.org/; for liability to coronary artery disease from the CARDIoGRAM consortium: http://www.cardiogramplusc4d.org/; and for liability to peripheral artery disease from dbGAP: https://www.ncbi.nlm.nih.gov/gap/. Approved dbGAP access to phs001672.v6.p1 was provided to B.F.V. (dbGAP project ID: 27398).

## FUNDING STATEMENT

VMW, ARC, NMD, GDS and TRG are members of the Medical Research Council Integrative Epidemiology Unit, which is supported by the Medical Research Council and the University of Bristol [MC_UU_00011/1, MC_UU_00011/4 and MC_UU_00011/6]. ARC is also supported by the University of Bristol British Heart Foundation Accelerator Award (AA/18/7/34219). NMD is also supported by a Norwegian Research Council [grant number 295989]. BFV is supported by the NIH/NIDDK (DK126194 and DK101478). M.S.U. is supported by NIDDK K23DK114551.

## CONFLICTS OF INTEREST STATEMENT

TRG receives funding from Biogen for research unrelated to this manuscript. The authors of this manuscript have no other conflicts of interest to declare.

## ETHICS APPROVAL STATEMENT

This research using UK Biobank was completed under Application Number 15825, which has been subject to ethics approval.

## CONTRIBUTION STATEMENT

VMW, BV, TRG, and SMD contributed to the design of the work. VMW performed the data analysis and drafted the article. All authors contributed to the data interpretation, critical revision of the article, and final approval for publication.

## ACKNOWLEDGEMENTS

Quality Control filtering of the UK Biobank data was conducted by R. Mitchell, G. Hemani, T. Dudding, Corbin, S. Harrison, L. Paternoster as described in the published protocol (doi: 10.5523/bris.1ovaau5sxunp2cv8rcy88688v). The MRC IEU UK Biobank GWAS pipeline was developed by B. Elsworth, R. Mitchell, C. Raistrick, L. Paternoster, G. Hemani, T. Gaunt (doi: 10.5523/bris.pnoat8cxo0u52p6ynfaekeigi).

The authors thank Million Veteran Program (MVP) staff, researchers, and volunteers, who have contributed to MVP, and especially participants who previously served their country in the military and now generously agreed to enroll in the study. (See https://www.research.va.gov/mvp/ for more details). The citation for MVP is Gaziano, J.M. et al. Million Veteran Program: A mega-biobank to study genetic influences on health and disease. J Clin Epidemiol 70, 214-23 (2016). This research is based on data from the Million Veteran Program, Office of Research and Development, Veterans Health Administration, and was supported by the Veterans Administration (VA) Cooperative Studies Program (CSP) award #G002.

## Notes

### Author Declarations

This manuscript used publicly available summary level data from previously published studies and so did not require specific IRB and/or ethics committee approval. Details of the IRB and/or ethics committee approval obtained for each contributing study can be found in their respective publications.

