## Supplementary Figure for "Separating the direct effects of risk factors for atherosclerotic cardiovascular disease from those mediated by type 2 diabetes"

### Separating the effects of risk factors from type 2 diabetes on coronary and peripheral artery disease

Supplementary Figures

Supplementary Figure 1: Flow chart showing selection of risk factors for the analysis

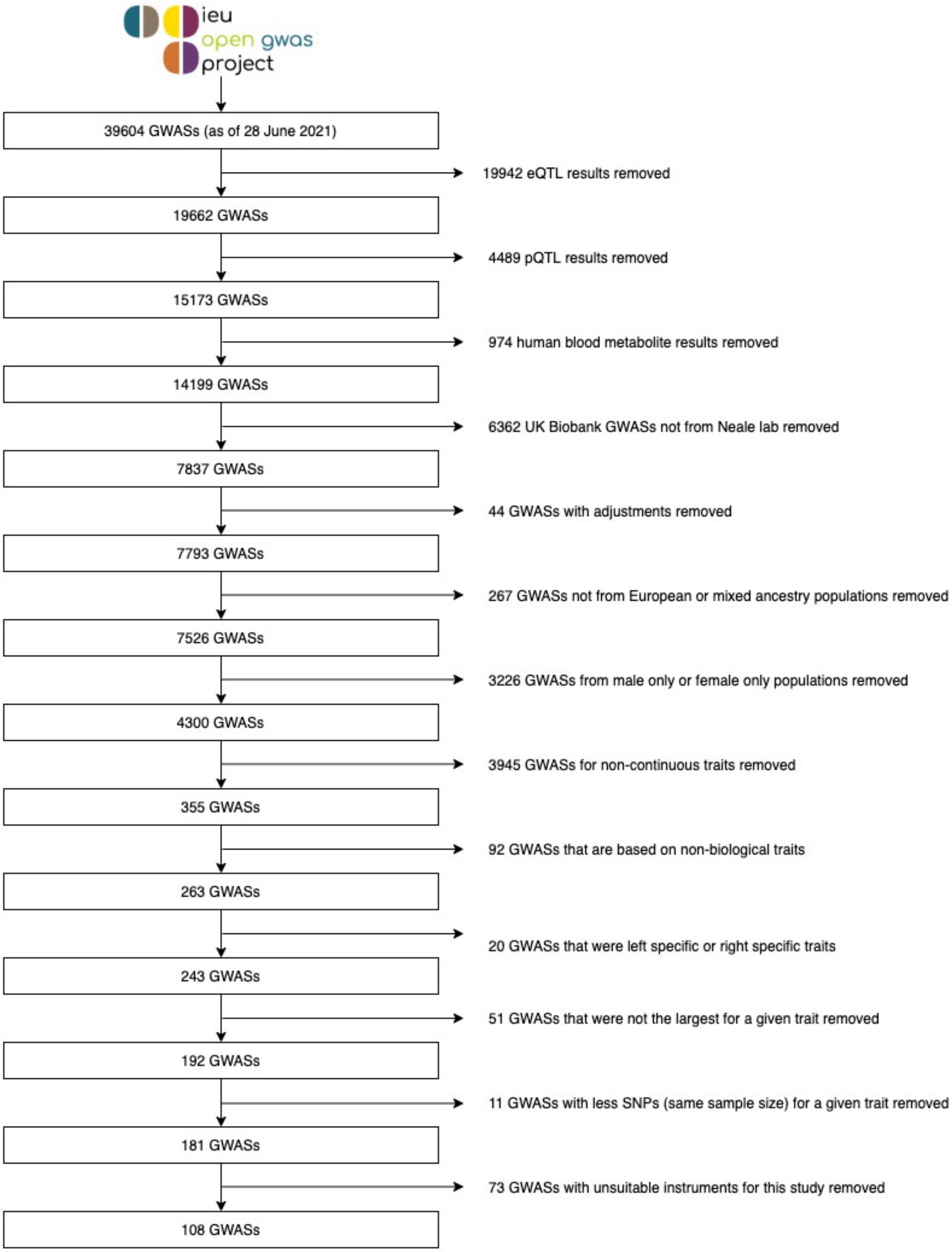

Supplementary Figure 2: Univariate Mendelian randomization estimates for the effect of the risk factors on liability to type 2 diabetes that meet the 5% FDR threshold (see Supplementary Table 2 for all estimates)

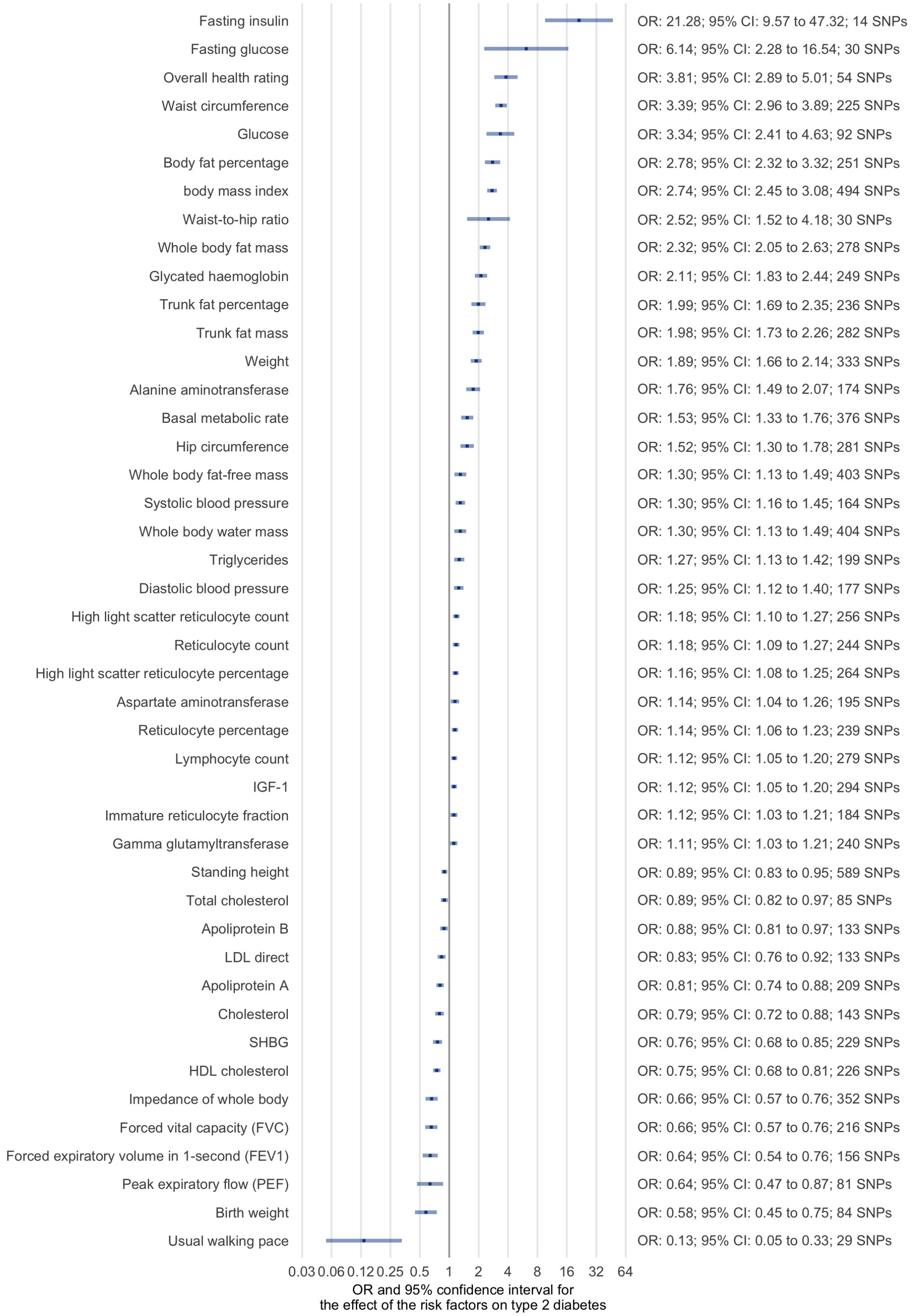

Supplementary Figure 3: Univariate Mendelian randomization estimates for the effect of liability to type 2 diabetes on the risk factors that meet the 5% FDR threshold (see Supplementary Table 2 for all estimates)

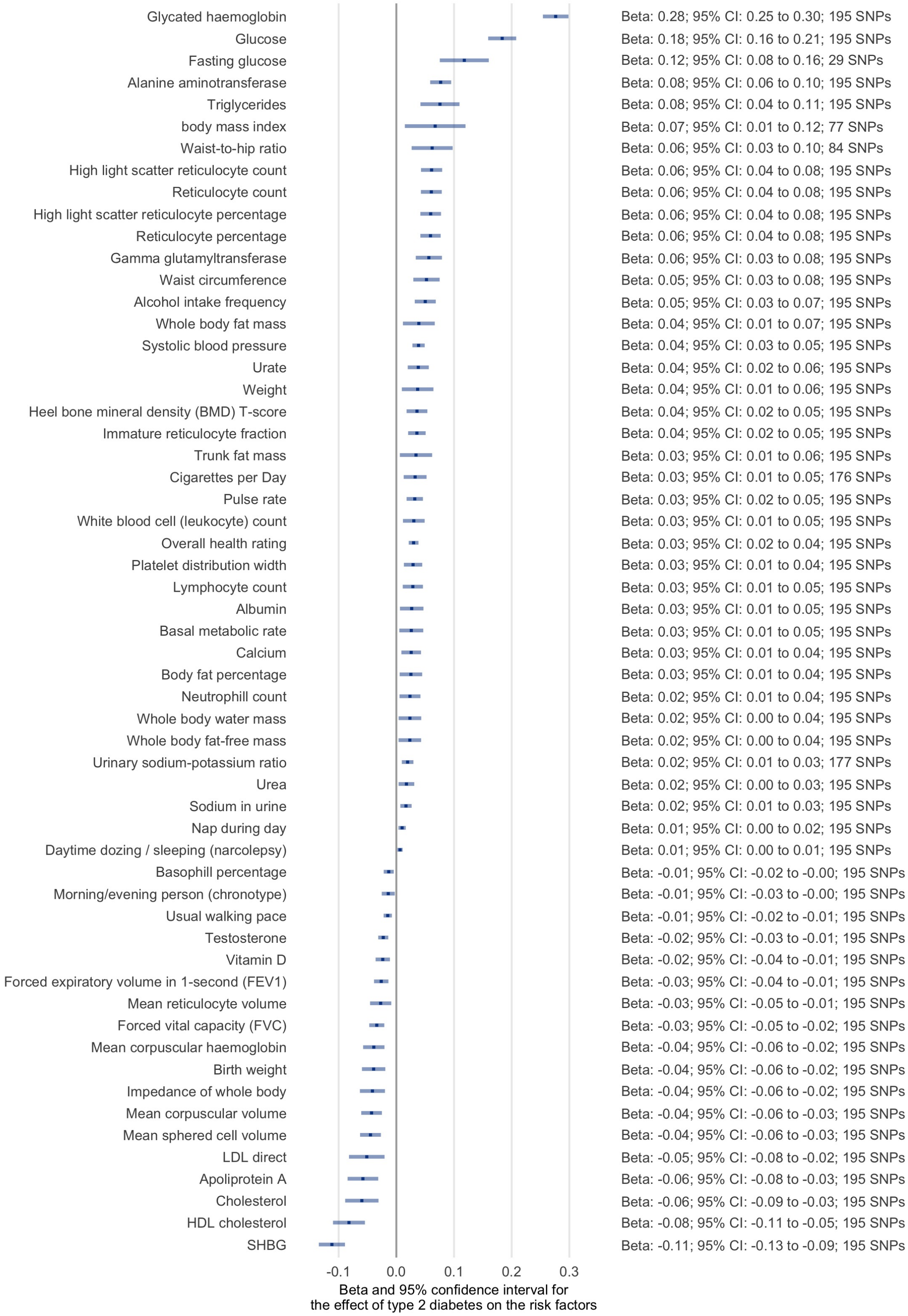

Supplementary Figure 4: Univariate Mendelian randomization estimates for the effect of the risk factors on liability to type 2 diabetes using alternative methods

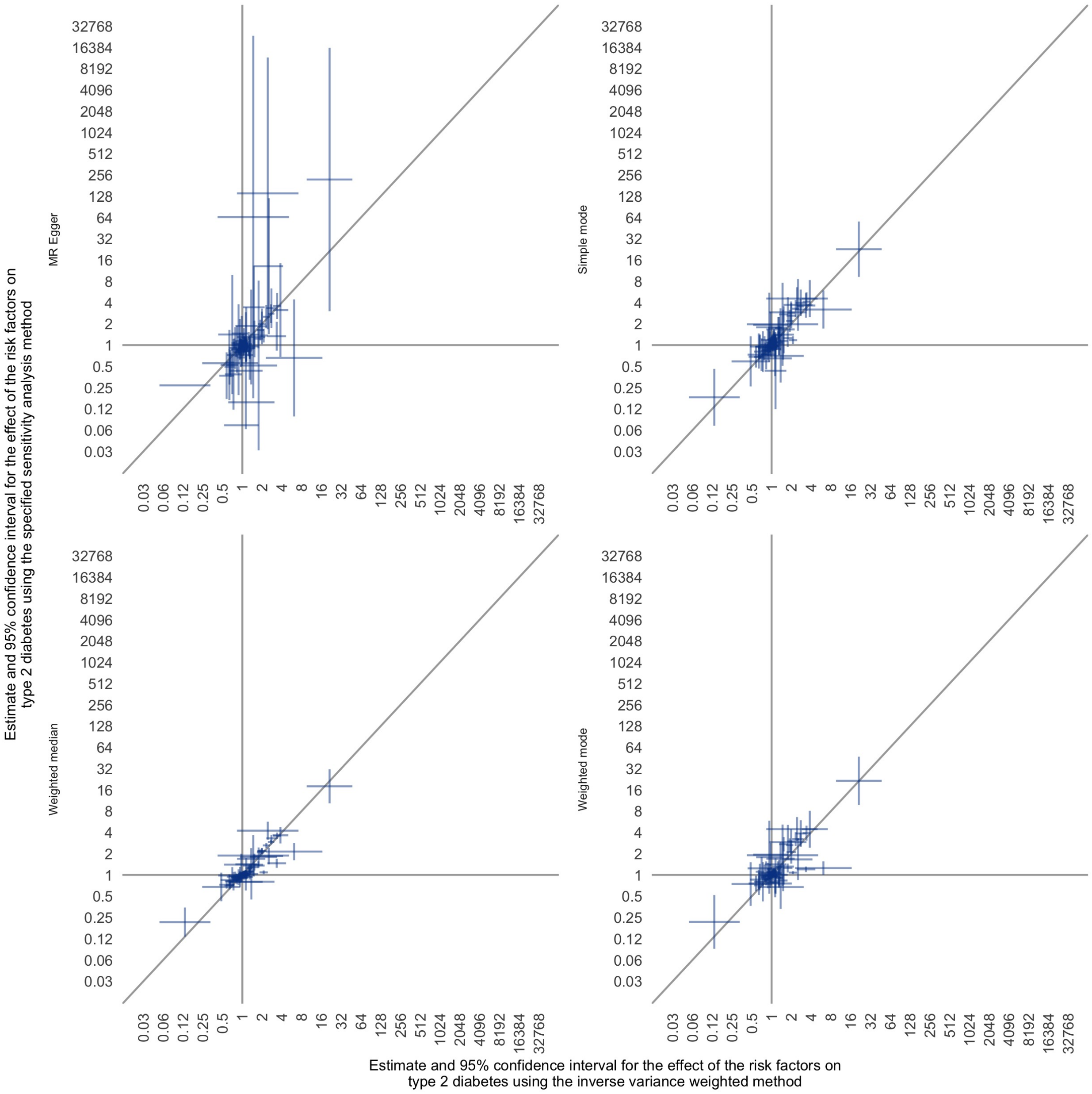

Supplementary Figure 5: Univariate Mendelian randomization estimates for the effect of liability to type 2 diabetes on the risk factors using alternative methods

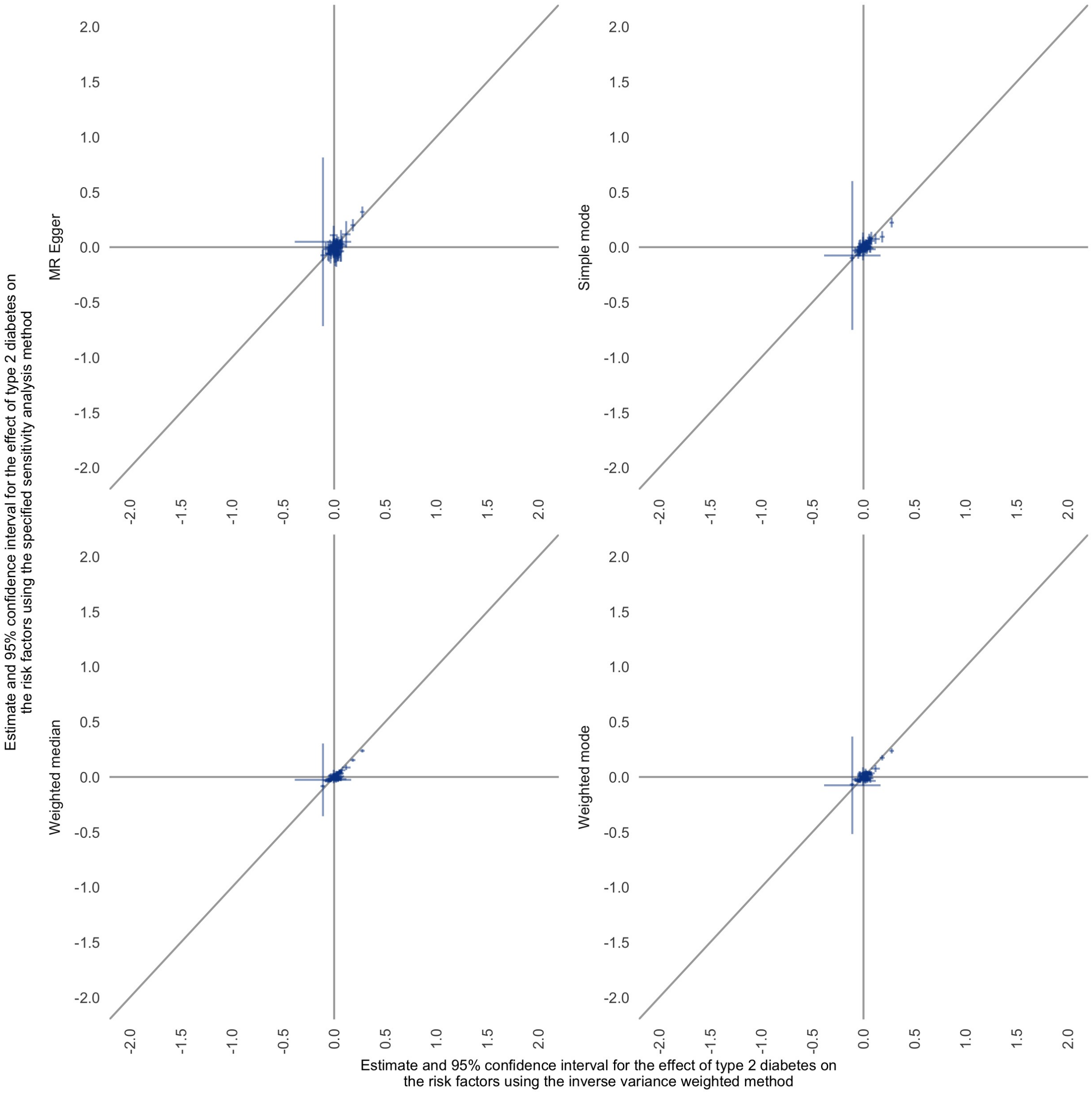

Supplementary Figure 6: Univariate Mendelian randomization estimates for the effect of the risk factors on liability to coronary artery disease that meet the 5% FDR threshold (see Supplementary Table 2 for all estimates)

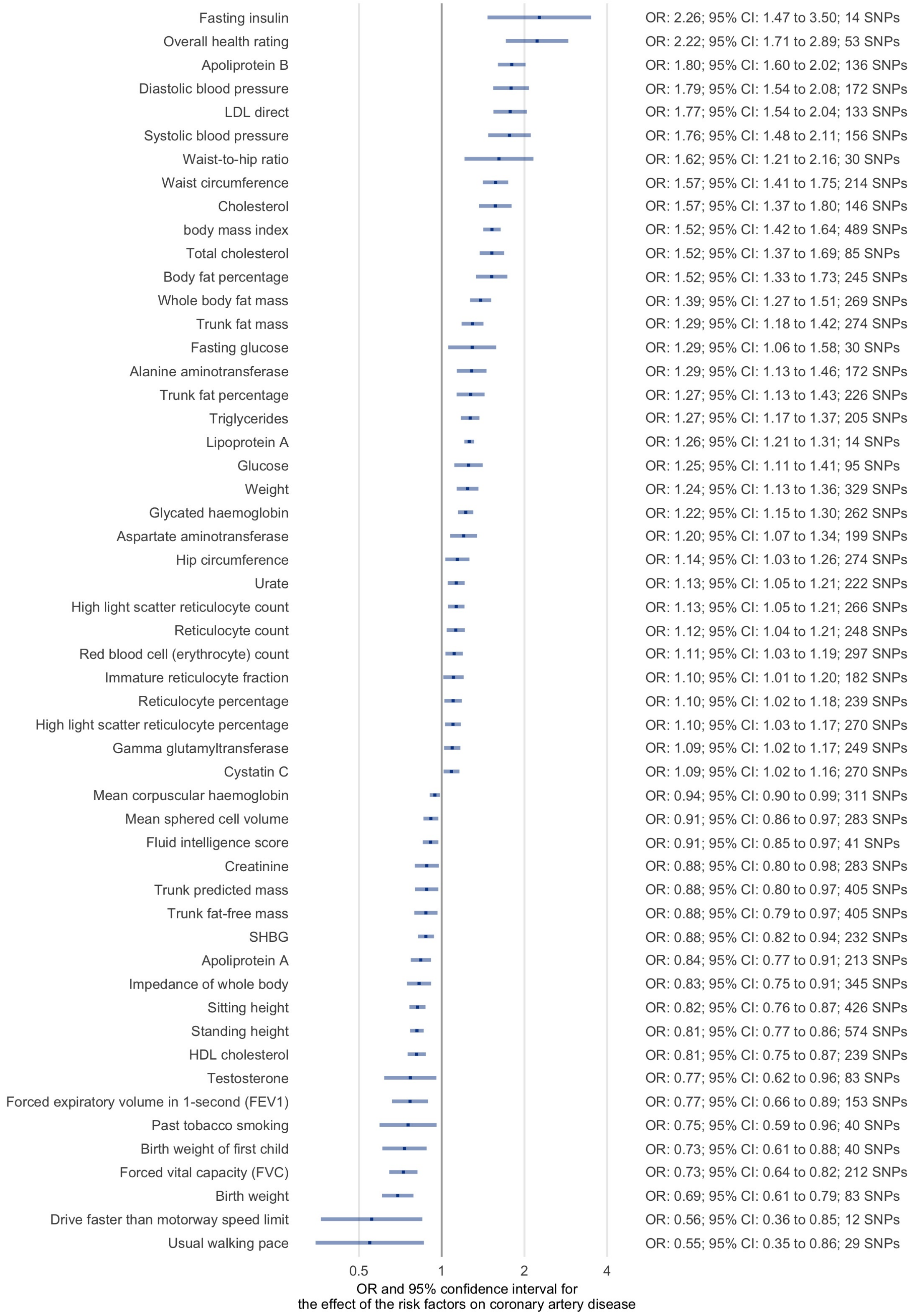

Supplementary Figure 7: Univariate Mendelian randomization estimates for the effect of the risk factors on liability to coronary artery disease using alternative methods

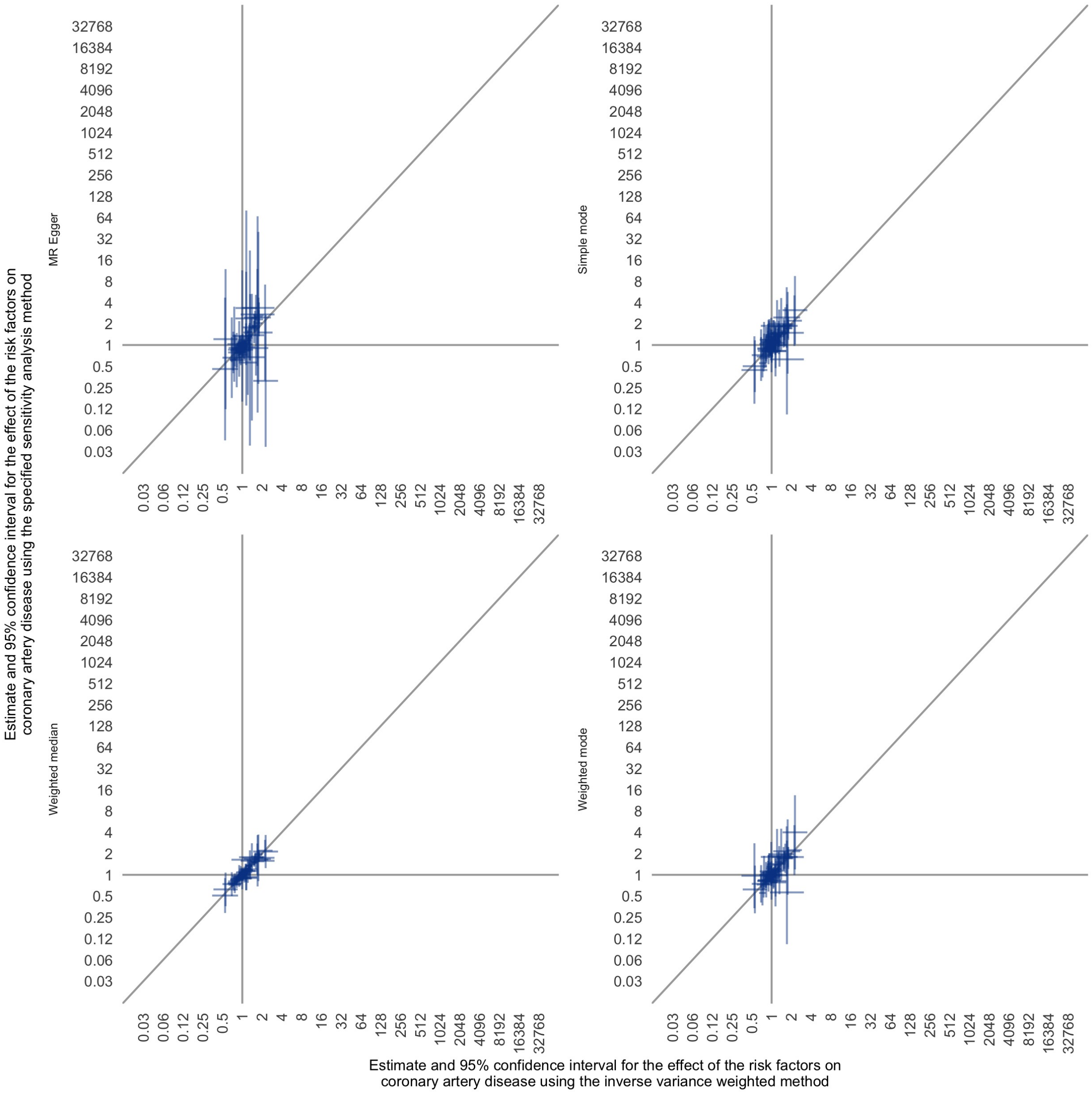

Supplementary Figure 8: Univariate Mendelian randomization estimates for the effect of the risk factors on liability to peripheral artery disease that meet the 5% FDR threshold (see Supplementary Table 2 for all estimates)

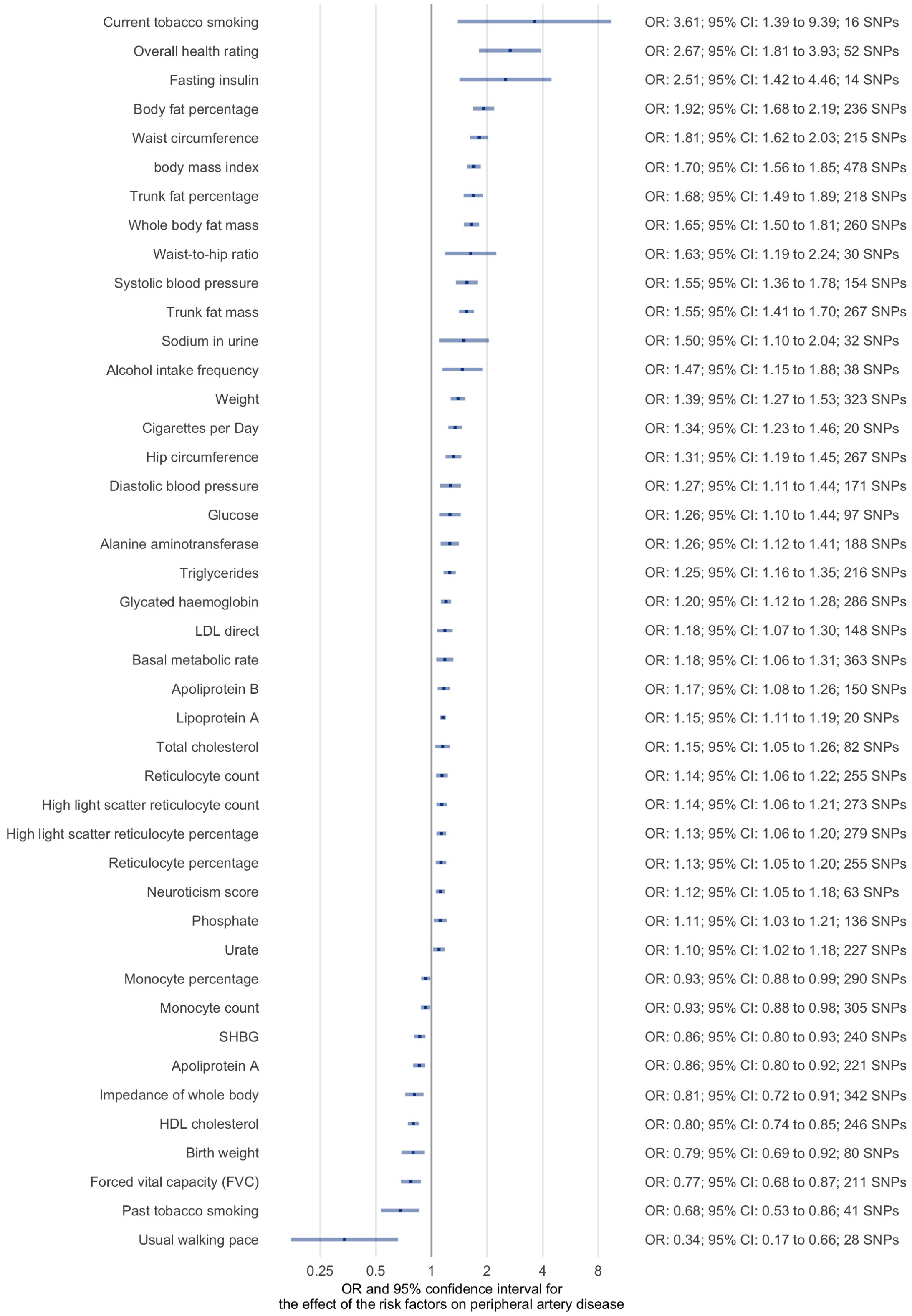

Supplementary Figure 9: Univariate Mendelian randomization estimates for the effect of the risk factors on liability to peripheral artery disease using alternative methods

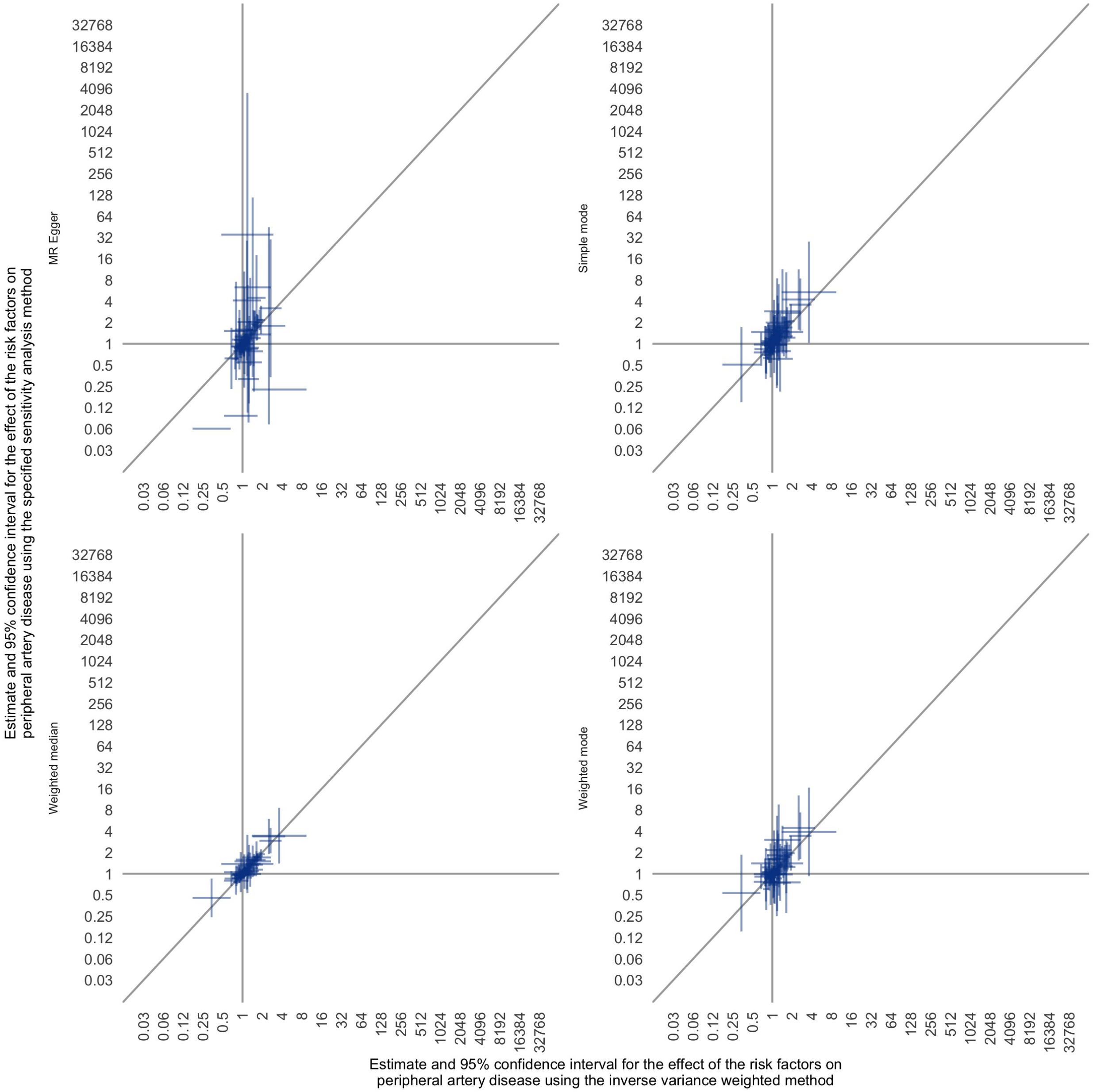

Supplementary Figure 10: Comparison plot illustrating the difference in all Mendelian randomization estimates when using a linear GWAS model for liability to type 2 diabetes versus a logistic GWAS model for liability to type 2 diabetes

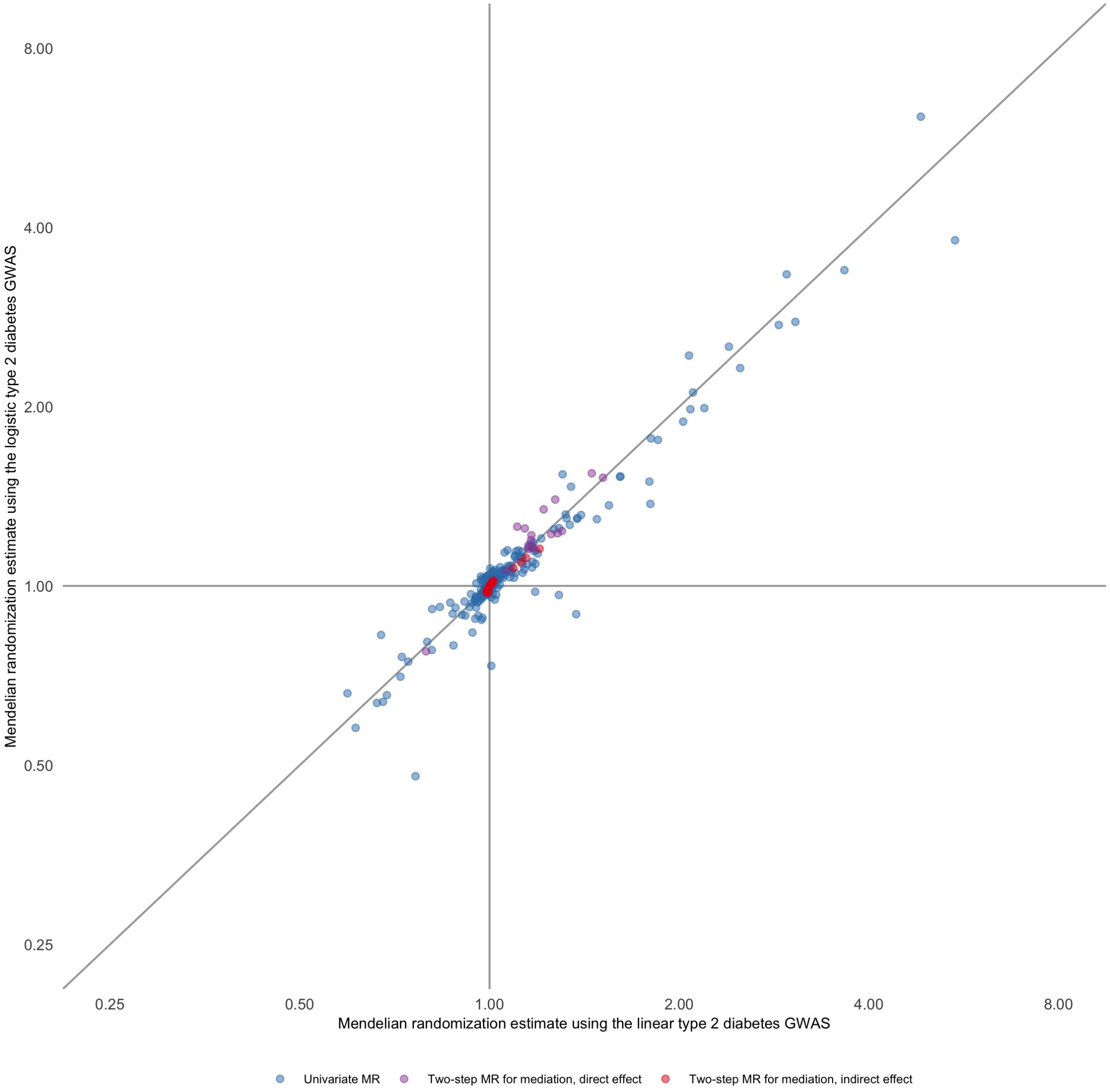
