## Supplementary Text for "Separating the direct effects of risk factors for atherosclerotic cardiovascular disease from those mediated by type 2 diabetes"

**Supplementary Text 1: Mendelian randomization analysis assumptions**

Mendelian randomization requires three assumptions: (1) the instrument must affect the exposure (relevance), (2) the instrument must not share any common causes with the outcome (independence), and (3) the instrument must only affect the outcome through the exposure (exclusion restriction). To obtain a point estimate, we must make a further assumption, such as monotonicity. This assumption states that the exposure is a monotonic (i.e. an always increasing or always decreasing) function of the instrument (monotonicity). Mendelian randomization performed using MR-Egger requires two alternative assumptions: the instrument strength independent of direct effect (INSIDE) assumption and the no measurement error (NOME) assumption. Finally, two-step Mendelian randomization for mediation assumes no interaction between the exposure and the mediator.

**Supplementary Text 2: Non-collapsibility of odds ratio**

The non-collapsibility of odds ratios can pose a problem when using summary statistics from logistic regression for binary mediators and outcomes in multivariable Mendelian randomization. To assess whether this is likely to have impacted our results, we performed a novel GWAS of liability to type 2 diabetes using a linear mixed model. Specifically, we performed a GWAS of type 2 diabetes using 24884 cases and 437996 controls from UK Biobank. The GWAS was conducted using the Medical Research Council Integrative Epidemiology Unit GWAS pipeline. Further details regarding the pipeline can be found here: https://doi.org/10.5523/bris.pnoat8cxo0u52p6ynfaekeigi. Type 2 diabetes was defined as a binary variable based on the presence of the ICD-10 code 'E11' as a main or secondary diagnosis in the hospital inpatient admissions data (UK Biobank data-field 41270). We used a BOLT-LMM model for the GWAS and adjusted for age, sex and chip. Individuals whose genetic sex did not match their reported gender; individuals with sex chromosome karyotypes putatively different from XX or XY; individuals who were outliers in heterozygosity and missing rates; and individuals with high levels of relatedness (3rd degree) to more than 200 other individuals in the biobank were excluded prior to the analysis. The GWAS is publicly available from the IEU OpenGWAS project (id: TBA). The code related to the GWAS can be found here: https://github.com/venexia/T2DLinearGWAS-UKB. We were then able to repeat our Mendelian randomization analyses, where there was no sample overlap with UK Biobank, using this GWAS and compare the results with our main analysis to assess the impact on our results.
